# Improving Heart Disease Probability Prediction Sensitivity with a Grow Network Model

**DOI:** 10.1101/2024.02.28.24303495

**Authors:** Simon Bin Akter, Rakibul Hasan, Sumya Akter, Md. Mahadi Hasan, Tanmoy Sarkar

## Abstract

The traditional approaches in heart disease prediction across a vast amount of data encountered a huge amount of class imbalances. Applying the conventional approaches that are available to resolve the class imbalances provides a low recall for the minority class or results in imbalance outcomes. A lightweight GrowNet-based architecture has been proposed that can obtain higher recall for the minority class using the Behavioral Risk Factor Surveillance System (BRFSS) 2022 dataset. A Synthetic Refinement Pipeline using Adaptive-TomekLinks has been employed to resolve the class imbalances. The proposed model has been tested in different versions of BRFSS datasets including BRFSS 2022, BRFSS 2021, and BRFSS 2020. The proposed model has obtained the highest specificity and sensitivity of 0.74 and 0.81 respectively across the BRFSS 2022 dataset. The proposed approach achieved an Area Under the Curve (AUC) of 0.8709. Additionally, applying explainable AI (XAI) to the proposed model has revealed the impacts of transitioning from smoking to e-cigarettes and chewing tobacco on heart disease.

## I. Introduction

Cardiovascular diseases are now recognized as a major global health problem, responsible for a significant number of deaths worldwide. Coronary artery disease, myocardial infarction, and congestive heart failure are among the most frequent and life-threatening of these conditions. Medical surveys include a wide range of health indicators, lifestyle habits, genetic variables, and patients’ past medical records [1] [2] [3]. These surveys have been used as an important source for cardiovascular research [2] [3] [4] [5]. Critical risk factors of heart disease have been possible to analyze through these data [4] [5]. However, these datasets need a high amount of preprocessing to produce useful results [1] [6]. To solve this problem, enhanced computational approaches are needed.

Machine learning and deep learning advancements in recent years have transformed the field of medical survey analysis [7] [8] [9]. Machine learning algorithms are effective in identifying patterns and predicting outcomes from medical survey data [4] [5]. Deep learning models, particularly Convolutional Neural Networks (CNNs) and Artificial Neural Networks (ANNs), have excelled in heart disease prediction using survey data, contributing to enhanced prediction accuracy [3] [10].

Many recognized studies utilizing artificial intelligence (AI) models showed high accuracy, nevertheless without additional information to confirm the dependability [2] [11]. Class imbalances are common among medical survey data [1] [6]. It is challenging to achieve high accuracy for the minority class while remaining well-balanced [1] [6] [12]. Sensitivity and specificity analyses are therefore vital for demonstrating such reliability and ensuring balanced outcomes. Prior studies that attempted such additional analyses found that the sensitivity and specificity were eventually unbalanced [1] [6] [12], resulting in biased results - an important problem that the proposed strategy in this work addresses. Prior studies that showed feature significance in predicting heart disease failed to provide precise information [11]. These analyses offered a summary of feature importance, without indicating the significance for each target class independently [11]. For this sort of study, finding the key variables that impact the prediction of certain target classes is essential for efficient model interpretation and decision-making [13]. Despite significant breakthroughs in the field of heart disease detection using machine learning and deep learning, there are still these identified challenges to be overcome [1] [6] [11] [14]. These issues underline the importance of this study in adding to the present body of knowledge and providing solutions to the difficulties outlined in the prior studies [1] [6] [10] [11] [14].

The present study for predicting heart disease employed a Synthetic Refinement Pipeline using Adaptive-TomekLinks to handle the class imbalances. A lightweight GrowNet-based architecture has been proposed that can obtain higher accuracy for the minority class while maintaining a balance. The interpretability of the proposed method has been determined using SHapley Additive exPlanations (SHAP) [15] [16], resulting in the identification of various risk variables associated with heart disease.

The key contributions and identified observations are summarized below.

- The proposed lightweight GrowNet model obtained high recall (0.81) for the minority class (heart disease) while maintaining a balanced recall (0.74) for the majority class (healthy) across heavily unbalanced data. This was a main challenge in heart disease prediction that many prior studies [1] [6] [12] struggled to ensure.
- A Synthetic Refinement Pipeline was proposed to handle the class imbalance which outperformed the other traditional approaches in balancing the outcomes using the proposed GrowNet model.
- The interpretability of the proposed GrowNet model, as measured by Shapley values [15], reveals significant risk indicators of heart disease. For example, the transition from cigarettes to chewing tobacco or e-cigarettes reduced the SHAP probability of heart disease.

In summary, this section addresses the necessity to improve heart disease prediction using medical survey data. The next section addresses the most relevant literature on the application of artificial intelligence (AI) in cardiovascular research using survey data. Following, the dataset part includes information about the overall number of samples and the distribution of target classes. Consecutively, the methodology section outlines the whole technical process of the proposed approach to reach the final outcomes. Later on, the feature description provides details for a list of relevant features chosen for this study based on prior studies. Sequentially, the preprocessing section includes data wrangling phases such as missing value handling, train-test splitting, label encoding, and balanced training using the Synthetic Refinement Pipeline. Furthermore, the next section describes the architectural details of the proposed Grownet model. The experiment and result analysis section next compares the findings obtained using various performance metrics for all of the applied models. Following, the interpretability analysis portion depicts the influence of each input characteristic on predicting heart disease using SHapley Additive ExPlanations (SHAP) values. Finally, the discussion section provides a complete summary of the study’s findings and observations.

## II. Literature review

This section reviews prior studies on the use of machine learning and deep learning to predict cardiovascular diseases, particularly heart disease. The objective is to identify gaps in the current body of knowledge and offer concise justifications for these limitations. Through a review of prior studies, the goal is to pinpoint areas in need of more study and development, which will ultimately lead to a more thorough and efficient method of comprehending and predicting heart-related diseases using artificial intelligence (AI) approaches.

A study conducted by Nasimov et al. [11] provided a methodology to reduce variations in the feature importance from the same model using different approaches for predicting chronic heart diseases. The initiative to show feature importance hueing the predictive classes wasn’t conducted in this study. This is essential to understand the impact of the input features on predictions. Accordingly, an accuracy of 0.743 was obtained from a weighted K-Nearest Neighbors (K-NN) model using the BRFSS 2021 dataset. The study [11] was conducted using a reduced and balanced dataset (27392 healthy and 23893 HD records). A similar study on heart disease risk prediction was conducted by Singh et al. [2] using the UCI Cleveland Heart Disease (HD) dataset. An accuracy of 0.87 was obtained using a K-Nearest Neighbors (K-NN) model. The study [2] was conducted using a balanced dataset with limited records (303 records) and the test set contained very limited data (51 healthy and 49 HD records). In the above studies [2] [11], there was no mention of any other important performance metrics other than accuracy to rely on the outcomes obtained by these studies. The test sets were limited and balanced in these studies [2] [11] so high accuracies are usual in these regards. To rely on these outcomes based on only accuracy is not enough as these accuracies can vary in terms of real-life scenarios (heavily unbalanced circumstances) [1] [6].

An effective approach for heart disease prediction was conducted by Bhatt et al. [10] using a dataset from Kaggle. The obtained accuracy, precision, recall, f1 score, and AUC were 0.8728, 0.887, 0.8485, 0.8671, and 0.95 respectively using a Multilayer Perceptron (MLP) model. The study [10] was conducted using a preprocessed dataset with limited records (57155 records) and the test set contained only 20% of the total data. Accordingly, a study on heart disease prediction was conducted by Pathan et al. [14] using two datasets from Kaggle including CVD and Framingham. For the CVD dataset, the obtained accuracy, f1 score, and AUC were 0.75, 0.74, and 0.74 respectively using a Support Vector Classifier (SVC) model. Further, for the Framingham dataset, the obtained accuracy, f1 score, and AUC were 0.72, 0.71, and 0.72 respectively using a Multilayer Perceptron (MLP) model. The study [14] was conducted using two reduced and balanced datasets (CVD: 548 healthy, 548 HD records and Framingham: 557 healthy, 557 HD records). A study to enhance heart disease prediction was conducted by Chandrasekhar et al. [5] employing an ensemble model (soft voting: RF, KNN, LR, GNB, GB, AdaBoost) using two datasets including UCI Cleveland Heart Disease (HD) and IEEE Dataport Heart Disease (HD). For the UCI Cleveland dataset, the obtained accuracy, precision, recall, f1 score, and AUC were 0.9344, 0.88, 0.84, 0.84, and 0.95 respectively. Further, for the IEEE Dataport dataset, the obtained accuracy, precision, recall, f1 score, and AUC were 0.95, 0.90, 0.89, 0.90, and 0.95 respectively. The study [5] was conducted using two limited and balanced datasets (UCI Cleveland: 138 healthy, 164 HD records and IEEE Dataport: 410 healthy, 508 HD records) and test sets were also very limited (UCI Cleveland: 27 healthy, 34 HD records and IEEE Dataport: 88 healthy, 96 HD records) accordingly. An interpretable approach for stroke prediction was conducted by Srinivasu et al. [13] using the Kaggle Stroke Prediction dataset. The obtained accuracy, precision, recall, f1 score, and AUC were 0.95, 1, 0.189, 0.038, and 0.659 respectively using an Artificial Neural Network (ANN) model. The study [13] was conducted using a limited and balanced dataset (4861 healthy, 249 HD records) and test sets were also very limited (972 healthy, 50 HD records) accordingly. Explainable AI (XAI) such as LIME [17] [18] was used to interpret the model, however, no initiative was conducted to show feature importance hueing the predictive classes. An approach to predict coronary heart disease and risk assessment was conducted by Huang et al. [12] using a collected dataset. The obtained accuracy, specificity, sensitivity, and AUC were 0.7896, 0.5113, 0.9386, and 0.8375 respectively using a Random Forest (RF) model. The study [12] was conducted using a preprocessed dataset with limited records (1502 records). A study for predicting cardiovascular heart disease was conducted by Srinivasan et al. [4] using the UCI Cleveland Heart Disease (HD) dataset. The obtained accuracy, specificity, sensitivity, precision, recall, f1 score, and AUC were 0.9878, 0.971, 0.9791, 0.9807, 0.9531, 0.9789, and 0.96 respectively using a Learning Vector Quantization (LVQ) model. In the above studies [4] [5] [10] [12] [13] [14] there was mention of much more important metrics rather than accuracy to rely on the outcomes obtained by these studies. However, these studies [4] [5] [10] [12] [13] [14] were evaluated across a limited and balanced number of records. These measures can vary in terms of heavily unbalanced scenarios which is more usual compared to real-life scenarios [1] [6]. Hence, it is not sufficient to rely on these measures as these were tested across very limited and balanced scenarios.

A comparative study to predict heart disease was conducted by Akkaya et al. [1] using the BRFSS 2020 dataset. The obtained accuracy, specificity, sensitivity, and AUC were 0.89, 0.94, 0.27, and 0.9487 respectively using an Extreme Gradient Boosting (XGB) model. The study [1] was conducted using a significant number of preprocessed data (280293 records) and the test set was also heavily unbalanced and large (51884 healthy and 4175 HD records). A similar study was conducted by Mamun et al. [6] to predict heart disease using the BRFSS 2020 dataset. The obtained accuracy, specificity, sensitivity, and AUC were 0.9157, 0.5261, 0.9232, and 0.84 respectively using a Logistic Regression (LR) model. The study [6] was conducted using a significant number of data (319795 records) and the test set was also large containing 20% of the preprocessed data. In the above studies [1] [6], there was mention of much more important metrics such as specificity and sensitivity to trust the outcomes obtained by these studies. Additionally, these studies [1] [6] were evaluated across a large and balanced number of records. However, the outcomes obtained from these studies [1] [6] were heavily unbalanced. In summary, some prior studies [2] [11] focused just on accuracy, which may not adequately represent a model’s performance. Simply having high accuracy does not ensure that the model accurately predicts both target classes [6]. Accordingly, some prior studies [3] [4] [5] [10] [12] [13] [14] employed various performance measures and received favorable evaluations, although these were conducted on small, balanced datasets. This may make the model look good, but real-world data is typically huge and imbalanced, thus the findings may be unreliable. On the other hand, a few prior studies [1] [6] used large imbalanced datasets, yet the results were likewise very unbalanced. As a result, there is a need for research that demonstrates how well a model operates across a vast amount of imbalanced scenarios while yet producing balanced results. The overview of the literature review is provided in **Table I**.

**TABLE I.** Summary of the literature review.

| Researcher | Year | Research Topic | Dataset | Modeling Technique | Accuracy |
| --- | --- | --- | --- | --- | --- |
| Bhatt et al. [10] | 2023 | Heart disease prediction | Kaggle HD | MLP | 0.8728 |
| Chandrasekhar et al. [5] | 2023 | Heart disease prediction | UCI HD, IEEE Dataport | Ensemble | 0.9344, 0.95 |
| Srinivasu et al. [13] | 2023 | Heart stroke prediction | Kaggle Stroke Prediction | ANN | 0.95 |
| Srinivasan et al. [4] | 2023 | Heart disease prediction | UCI HD | LVQ | 0.9878 |
| Akkaya et al. [1] | 2022 | Heart disease prediction | BRFSS 2020 | XGB | 0.89 |
| Mamun et al. [6] | 2022 | Heart disease prediction | BRFSS 2020 | LR | 0.9157 |
| Nasimov et al. [11] | 2022 | Heart disease prediction | BRFSS 2021 | weighted K-NN | 0.743 |
| Pathan et al. [14] | 2022 | Heart disease prediction | CVD, Framingham | SVC, MLP | 0.75, 0.72 |
| Huang et al. [12] | 2022 | Heart disease prediction | Collected | RF | 0.7896 |
| Singh et al. [2] | 2020 | Heart disease prediction | UCI HD | KNN | 0.87 |

## III. Dataset

The Behavioural Risk Factor Surveillance System (BRFSS) 2022 dataset [19] was employed in this study, which is a comprehensive collection of health-related information collected from surveys conducted throughout the United States. The dataset initially has 326 features with 445132 number of records. A careful review of the data reveals that the data distributions of healthy and heart disease cases in the target column are around 93.04% and 5.96%, indicating a continuing predominance of healthy instances. The dataset contains a wide range of health and lifestyle-related factors that are used to train the heart disease probability prediction model. The previous two versions of Behavioural Risk Factor Surveillance System datasets including BRFSS 2020 [19] and BRFSS 2021 [19] are also used in this study for testing the proposed GrowNet model.

## IV. Methodology

The phases to predict heart disease probability from heavily unbalanced survey data are discussed in this section. Obtaining a balanced recall for both predictive classes (healthy and HD) was a real challenge for this study. The first section describes the procedures used to acquire and prepare the data and includes preprocessing and data collection. The description of data splitting and methods for addressing class imbalance are provided next. The selection of an appropriate model is then discussed. The experiment, analysis of the results, and an interpretability analysis are finally discussed.

The technical steps to prepare the proposed heart disease probability prediction model are shown in **Fig 1**. Firstly, 12 features (6 binary and 6 multi-class) were selected from the dataset that contains health records, health habits, and demographic information of individuals. The features are selected based on the prior studies on heart disease [1] [2] [3] [4] [5] [6]. Specifically, the features that were mostly used in previous studies to predict cardiovascular diseases. Initially, the data contained a significant amount of missing inputs as the data was collected by telephone surveys. Besides, many participants refused to provide information, these were labeled as refused input in the data. Hence, data was preprocessed by cleaning all missing and refused inputs. Accordingly, data was split into separate train and test sets. Additionally, the data distribution for each category inside the selected features was heavily imbalanced. To provide stabilized training, multiple approaches were applied to handle the class imbalance. Multiple approaches such as Random Oversampling (ROS), Random Undersampling (RUS), Synthetic Minority Over-sampling Technique (SMOTE), Adaptive Synthetic Sampling Approach for Imbalanced Learning (ADASYN), Tomek-Links, Edited Nearest Neighbors (ENN), SMOTE Edited Nearest Neighbors (SMOTEENN), Near-Miss, SMOTE Undersampling, and Cost-Sensitive Learning (CSL) were applied to compare the capability of the proposed Synthetic Refinement Pipeline (SRP). The Synthetic Refinement Pipeline handled the class imbalance in a more optimized way compared to other approaches. Sequentially, the target column was encoded to set the prediction labels. Then multiple models such as Decision Tree (DT), Adaptive Boosting (AdaBoost), Multilayer Perceptron (MLP), Random Forest (RF), Artificial Neural Network (ANN), Convolutional Neural Network, Gaussian Naïve Bayes (GNB), Random Undersampling (RUSBoost), and GrowNet were trained using the prepared data to predict the heart disease cases. Then, the applied models were evaluated across different performance matrices. The proposed GrowNet model obtained high recall for the minority class (heart disease) while maintaining a balanced recall for the majority class (healthy) using the Synthetic Refinement Pipeline. Lastly, an interpretability analysis was conducted to explain the proposed GrowNet model’s hidden patterns. This demonstrates multiple risk factors for heart disease using the SHapley Additive exPlanations.

**Fig. 1.**
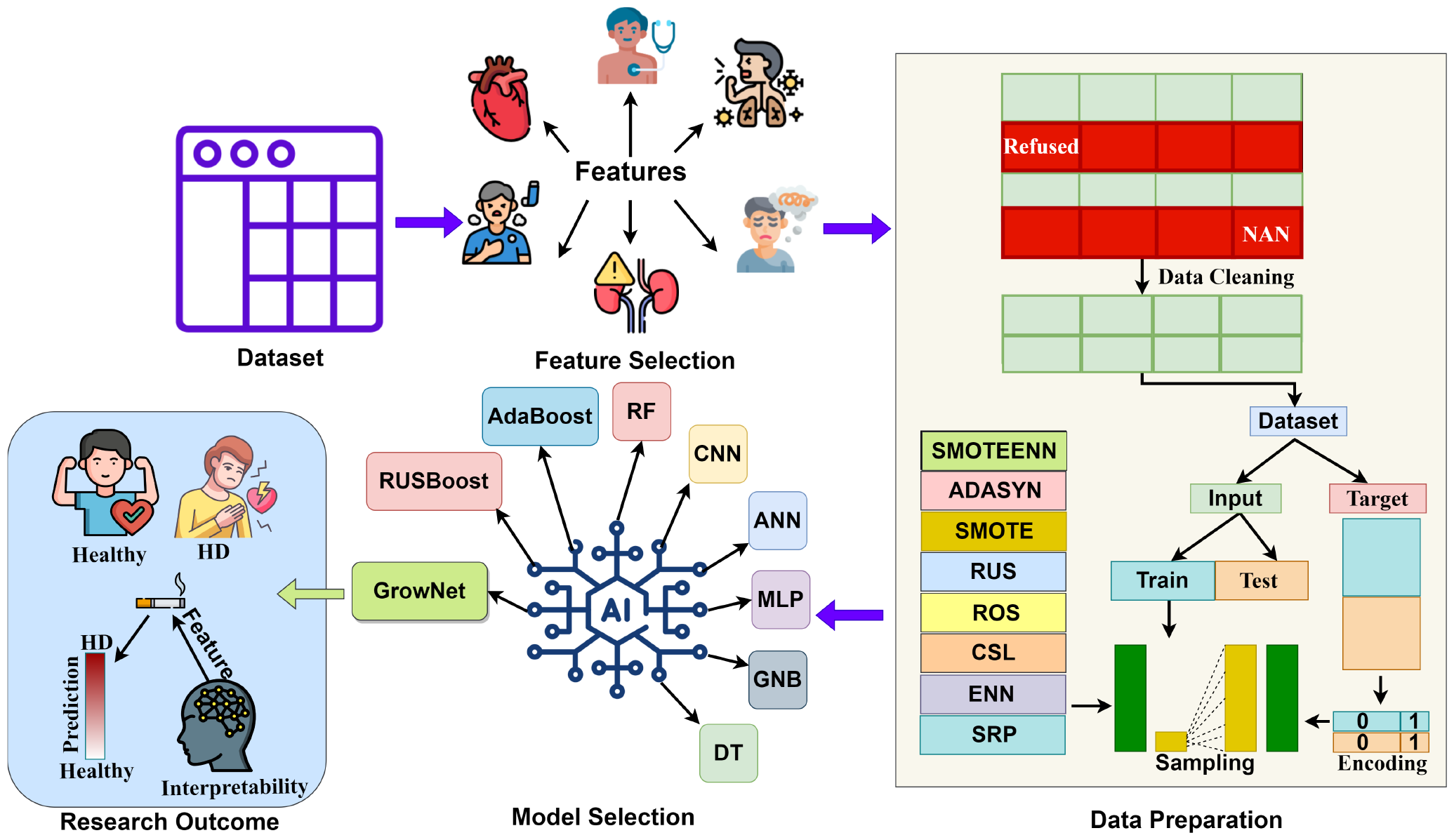
The complete workflow of the heart disease prediction.

## V. Feature description

Only 13 features are selected for this study out of 326 features from the dataset based on the prior studies [20] [21] [22]. These features are mostly covered medical records of the individuals such as histories of heart attacks, strokes, asthma, bronchitis, renal issues, depressive disorders, and Body Mass Index (BMI). Besides, It also covers a number of aspects of health-related behaviors, such as the status of smoking, the use of electronic cigarettes or other vaping devices, and the use of chewing tobacco, snuff, or snus. Additionally, it also contains an individual’s age, which is a demographic aspect.

The data distributions for each category inside the input features are discussed in **Table II**. The features including heart attack, depressive disorder, chronic bronchitis, stroke, renal issues, and asthma have binary categories. Accordingly, the features including age groups, diabetes, BMI, smoking status, tobacco usage, and e-cigarettes have multiple categories. The categorization inside these features seemed to be heavily unbalanced.

**TABLE II.** Data distribution of the input features.

| Feature | Frequency |
| --- | --- |
| Heart attack | 1 (yes: 25108), 2 (no: 416959) |
| Depressive disorder | 1 (yes: 91410), 2 (no: 350910) |
| Chronic bronchitis | 1 (yes: 35656), 2 (no: 407257) |
| Stroke | 1 (yes: 19239), 2 (no: 424336) |
| Renal issues | 1 (yes: 19239), 2 (no: 424336) |
| Asthma | 1 (yes: 20315), 2 (no: 422891) |
| Diabetes | 1 (yes: 61158), 2 (gestational: 3836), 3(no: 368722), 4(pre-diabetes: 10329) |
| Age groups | 1 (18 to 24: 26943), 2 (25 to 34: 47840), 3 (35 to 44: 59174), 4 (45 to 54: 66984), 5 (55 to 64: 82740), 6(65 or older: 161451) |
| BMI | 1 (BMI < 1850: 26943), 2 (1850 <= BMI < 2500: 47840), 3 (2500 <= BMI < 3000: 59174), 4 (3000 <= BMI: 66984) |
| Smoking status | 1 (regular: 36003), 2 (irregular: 13938), 3 (former: 113774), 4 (never: 245955) |
| Tobacco usage | 1 (regular: 7333), 2 (irregular: 5358), 3 (no: 398616) |
| E-cigarette | 1 (never: 311988), 2 (regular: 10382), 3 (irregular: 11734), 4 (former: 75368) |

## VI. Preprocessing

As the survey was carried out via telephone, it contained a significant number of missing values. The dataset contains 62241 rows with missing values. Besides, many respondents to the survey refused to answer questions or said they were unsure of the precise responses at the time. These circumstances are denoted with specific input categories in the dataset. Accordingly, 57588 rows are identified as these reused inputs. After cleaning these missing and refused inputs, the dataset now contains 367320 clean records. Then, the dataset is divided into two sections including input and target. Input contains 12 features and the target contains the feature that has heart disease information. The target column is encoded into two categories such as class 0 represents healthy cases and class 1 represents heart disease cases. Then the two sections input and target are divided with a split ratio of 51.22% for training and 48.78% for testing. This unusual distribution was chosen to obtain acceptable outcomes from heavily unbalanced data. There are initially 172472 records of healthy cases and 15662 records of heart disease cases in the train set. Similarly, the test set included 172473 records of healthy cases and 6713 records of heart disease cases. After balancing the train set using the Synthetic Refinement Pipeline, the train set now contains 172472 records of healthy cases and 173404 records of heart disease. Accordingly, the proposed model is tested on the BRFSS 2020 dataset that contains 46640 records (healthy: 44665 records, heart disease: 1975 records) after preprocessing. Similarly, the proposed model is also tested on the BRFSS 2021 dataset that contains 369767 records (healthy: 350042 records, heart disease: 19725 records) after preprocessing.

The equations for the Synthetic Refinement Pipeline:

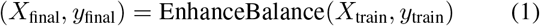

Here, operator EnhanceBalance uses a dynamic mix of oversampling and undersampling to improve the balance across classes.

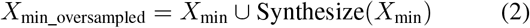

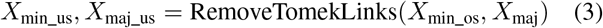

Here, ADASYN oversampling (os) procedure is Synthesize, while the Tomek Links undersampling (us) process is RemoveTomekLinks.

The Synthetic Refinement Pipeline is like a makeover for the dataset to enhance the balance. To address the class imbalance, it adds more variations to the minority class (heart disease) by creating diverse copies of these, balancing the predictive classes. Next, it removes unnecessary connections between the newly added data and the majority (healthy cases), creating a harmonious distribution of data. This dynamic process of adding and adjusting records, done through the EnhanceBalance method, ensures that the final train set has a better mix of both predictive classes (healthy and HD), making it fair and ready for training in artificial intelligence (AI) models.

## VII. Model architecture

A custom GrowNet was proposed to predict heart disease from heavily unbalanced survey data. GrowNet is a method for improving neural networks by progressively adding and refining neurons during training, increasing the model’s capacity to learn and predict accurately shown in **Fig 2**.

**Fig. 2.**
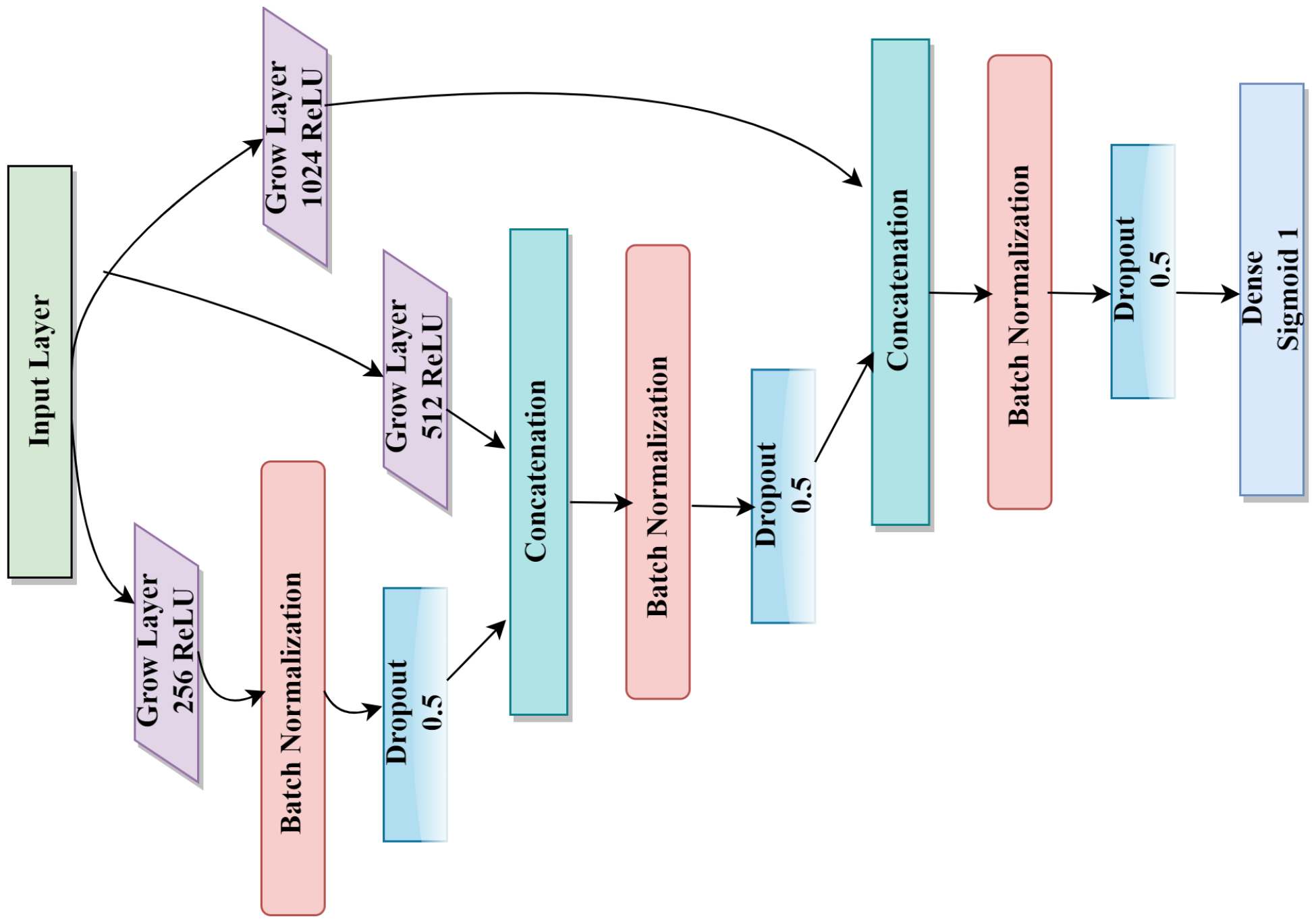
Architecture of the proposed GrowNet model.

The equations for the proposed GrowNet model:

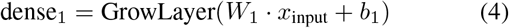

This equation reflects the initial dense layer’s (dense_1_) output in the GrowNet tabular model. The input is multiplied by the weight matrix (*W*_1_), and the output is fed into the GrowLayer activation function with bias (*b*_1_).

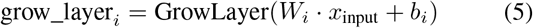

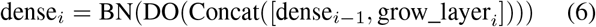

These equations explain the extra GrowNet layers (grow layer*i* and dense_*i*_) where *i* vary from two to three. The grow layer is computed similarly to the initial dense layer, while the dense layer incorporates batch normalization (BN), dropout (DO), and concatenation (Concat) with the preceding dense layer.

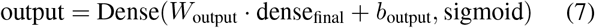

This equation corresponds to the GrowNet tabular model’s output layer (output). It employs a final dense layer with a sigmoid activation function to generate the model’s prediction.

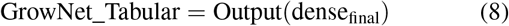

This equation explains the general structure of the GrowNet tabular model, with the ultimate output coming from the last dense layer.

The input layer of the Grow Neural Network design receives data in a given format. Employing the ReLU activation function, the GrowLayer is the first dense layer with 256 units. To standardize inputs, batch normalization is used, and dropout at a rate of 0.5 is used to avoid overfitting. There are two more GrowNet layers in the network, each of which doubles the total number of units. GrowLayer, batch normalization, and dropout form these layers. Every GrowLayer’s output is concatenated with the dense layer previous to it. Lastly, a dense layer with one unit and a sigmoid activation function forms the output layer, which is appropriate for applications involving binary classification. The architecture uses regularisation techniques like batch normalization and dropout to prevent overfitting and attempt to capture complicated patterns in the input data.

## VIII. Experiment and result analysis

A wide range of performance matrices [26] are used to analyze and compare the ability of the proposed prediction model. The balanced dataset refined through the Synthetic Refinement Pipeline was used in this section. First, training and validation metrics of the proposed GrowNet model, such as accuracy and loss, are measured at the epoch level. Further, the specificity and sensitivity of all the applied models are computed from datasets that are both unbalanced and balanced to measure the progress. The Area Under the Receiver Operating Characteristic (AUC) curves for all the applied models are assessed in the next section. Finally, the confusion matrix for the proposed GrowNet model was determined. Besides, confusion matrices for different models that outperformed other models using a traditional approach to address class imbalance are determined. This will provide an overview of the significance of the proposed sampling pipeline (Synthetic Refinement) in comparison to alternative strategies to handle class imbalance. Additionally, the confusion matrices for the proposed GrowNet model when tested on BRFSS 2020 and BRFSS 2021 datasets, are also figured.

Equations to compute the specificity [26] and sensitivity [27]:

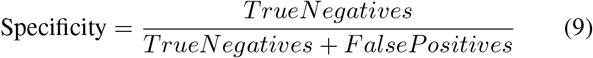

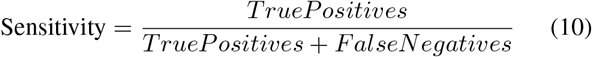

Specificity indicates how well a model correctly identifies those who don’t have a probability of heart disease, and sensitivity shows how well a model correctly identifies those who have a probability of heart disease. The obtained specificity and sensitivity across different approaches are shown in **Table III**. When trained on raw data, a boosting model (Random Undersampling Boosting) obtained balanced specificity and sensitivity. However, a Grow neural network-based model was proposed which obtained the most optimal specificity and sensitivity when trained on a stabilized dataset (balanced using a Synthetic Refinement Pipeline). The average performance of Random Undersampling Boosting (RUSBoost) initially looked better than the proposed GrowNet. However, the main intention of this study is to obtain better recall for the minority class while maintaining a balanced recall for the majority class. The proposed GrowNet model obtained a significantly higher recall for the heart disease class (minority class) than the RUSBoost and also maintained a balanced recall for the healthy class (majority class). A few models such as Adaptive Boosting (AdaBoost), Multilayer Perceptron (MLP), Random Forest (RF), and Artificial Neural Network (ANN) obtained higher recall for the heart disease class than the proposed GrowNet but also performed very poorly in predicting the healthy class with obtained recall below 70%. Hence, outcomes obtained from the proposed GrowNet were more optimized in balancing the recall for both classes prioritizing the minority class.

**TABLE III.** Specificity and sensitivity from raw and balanced data (stabilized using Synthetic Refinement Pipeline).

| Approaches | Training on Raw Data |  | Balanced Training |  |
| --- | --- | --- | --- | --- |
|  | Specificity | Sensitivity | Specificity | Sensitivity |
| DT | 0.98 | 0.35 | 0.75 | 0.68 |
| AdaBoost | 0.99 | 0.26 | 0.69 | 0.84 |
| MLP | 0.98 | 0.42 | 0.68 | 0.85 |
| RF | 1.00 | 0.00 | 0.66 | 0.86 |
| ANN | 0.98 | 0.43 | 0.64 | 0.88 |
| CNN | 0.98 | 0.42 | 0.76 | 0.76 |
| GNB | 0.89 | 0.65 | 0.84 | 0.70 |
| RUSBoost | 0.84 | 0.71 | 0.81 | 0.73 |
| **GrowNet** | **0.98** | **0.41** | **0.74** | **0.81** |

The training and validation phases are shown in **Fig 3**. The training phase went smoothly as the training dataset was balanced using a Synthetic Refinement Pipeline. However, few spikes were seen for validation losses and accuracies as there was a huge imbalance between the prediction classes in the test dataset. The highest validation accuracy of over 0.86 was obtained at epoch 5.

**Fig. 3.**
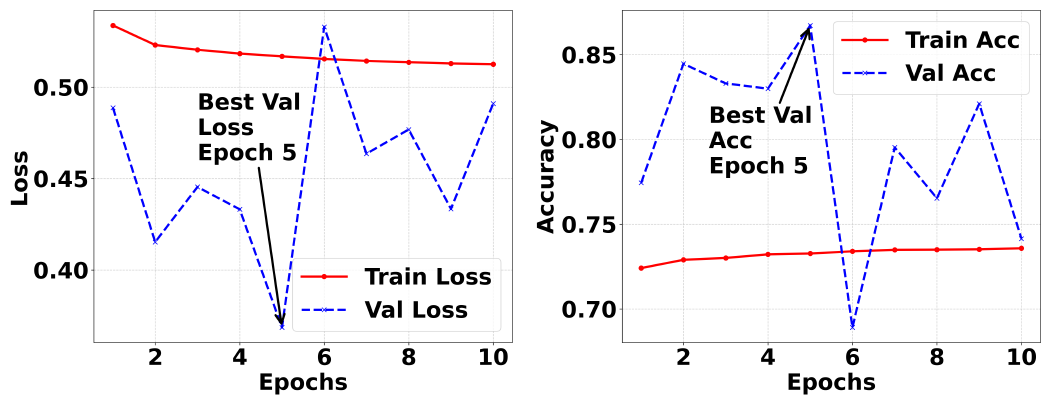
Training and validation phases within each epoch.

The Area Under a Receiver Operating Characteristic (ROC) curve provided in **Fig 4** depicts a model’s ability to distinguish between two predictive classes (healthy and HD cases). A higher AUC score indicates better overall performance in heart disease prediction. The AUC curve for GrowNet rises from the bottom-left corner which indicates higher true positive rates and lower false positive rates. This demonstrates that GrowNet is better at distinguishing between predictive classes (healthy and HD). Besides, the proposed grow network-based model outperformed the other applied models, displaying the highest AUC scores above 0.87.

**Fig. 4.**
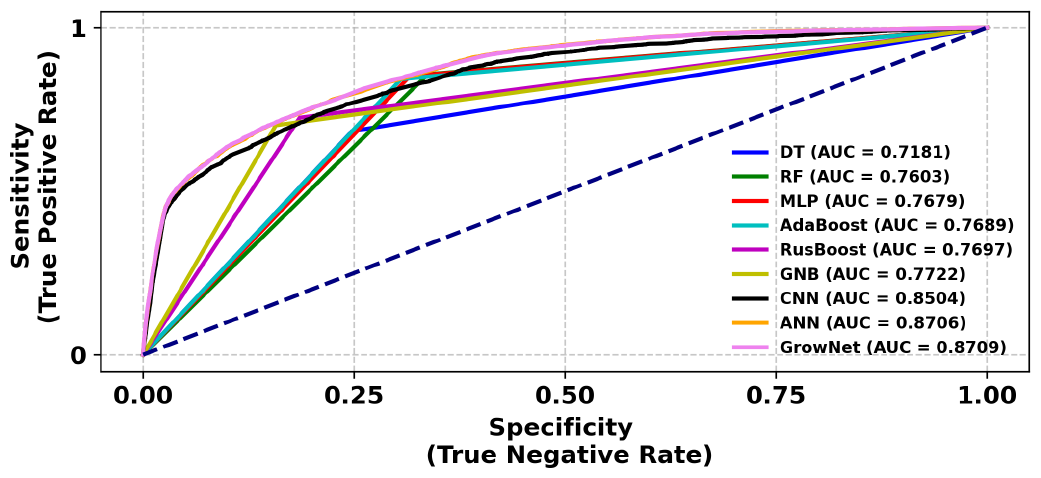
AUC curves for all the applied models.

The normalized confusion matrices in **Fig 5** depicts the proportion of correct predictions for each predictive class in the heart disease prediction. All the models are evaluated using different approaches for balancing the class imbalance such as Random Oversampling (ROS), Random Undersampling (RUS) [14], Synthetic Minority Oversampling Technique (SMOTE) [1], Adaptive Synthetic Sampling Approach for Imbalanced Learning (ADASYN) [13], Tomek-Links [1], Edited Nearest Neighbors (ENN) [13], SMOTE Edited Nearest Neighbors (SMOTEENN) [13], Near-Miss [13], SMOTE Undersampling [1], Cost-Sensitive Learning (CSL) [29], and proposed Synthetic Refinement Pipeline (SRP). For each sampling approach, the optimal confusion matrix achieved through a particular model is shown in this section. The proposed GrowNet model, trained on balanced data using the Synthetic Refinement Pipeline (SRP), obtained the most optimized recall in identifying heart disease cases across the BRFSS2022 dataset while maintaining a balance with the recall of healthy cases. The Convolutional Neural Network (CNN) model using cost-sensitive learning (CSL) obtained 92% recall for the heart disease class but obtained a very low around 58% recall for the healthy class. This approach obtained better recall for the heart disease class than the proposed GrowNet but was still not optimized for its significantly poor performance in predicting the healthy class. Besides, other applied approaches obtained less recall for the heart disease class compared to the proposed GrowNet’s recall in this regard. The main focus of this study is the minority class which is the heart disease class. Hence, the proposed GrowNet model performed in a more optimized way than other models. Additionally, the proposed model also obtained optimal recalls for heart disease cases across the BRFSS2020, and BRFSS2021 datasets, while also balancing the recall for healthy cases.

**Fig. 5.**
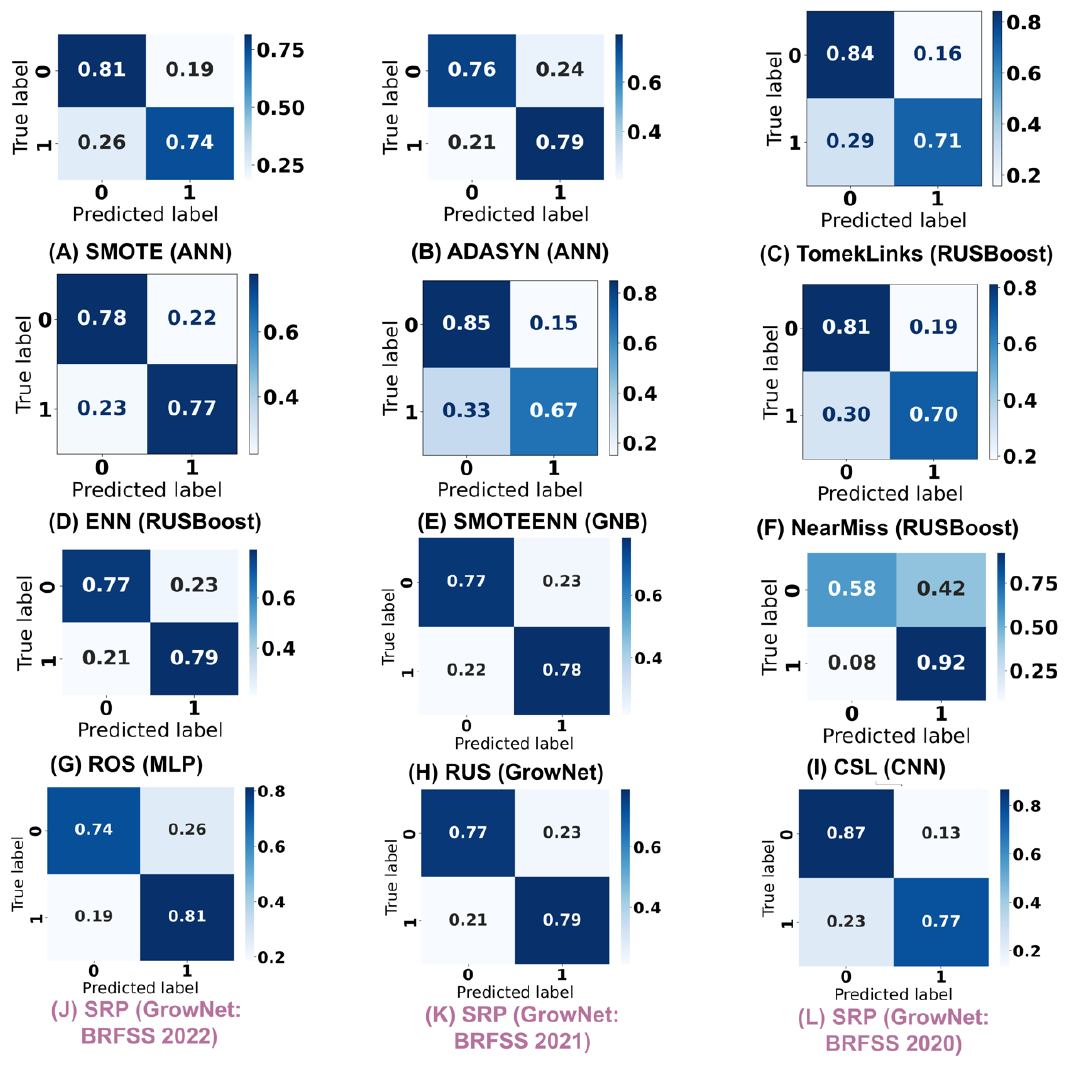
Confusion matrices for different sampling approaches. For any sampling strategy, the ideal confusion metric mentioning the model is shown here.

### A. Explainability

The sensitivity of the proposed model towards each input feature is analyzed here using SHapley Additive exPlanations (SHAP) [23] [24] [25]. First, the impact of all the input features is measured using the SHAP summary plot. This demonstrates the impact of each input category inside selected features hueing the predictive classes (healthy and HD). However, the features that have multiple categories inside are thoroughly discussed next for clear observations using the SHAP dependence plot. Next, the impacts of smoking, chewing tobacco, and e-cigarettes on heart disease are measured using the SHAP values.

The effects of the input features are shown in **Fig 6**. Individuals who have a history of heart attack, diabetes, depressive disorder, chronic bronchitis, stroke, renal issues, and asthma have a significant impact on the prediction of heart disease cases. Further, age groups and BMI categories have a good influence on heart disease prediction. Accordingly, health habits including smoking status, tobacco usage, and e-cigarettes have also a significant impact on the prediction of heart disease. The influence of each category for the binary columns is clearly visible. However, detailed analyses are required to understand the influence of the categories in multiclass columns which will be discussed in the next section.

**Fig. 6.**
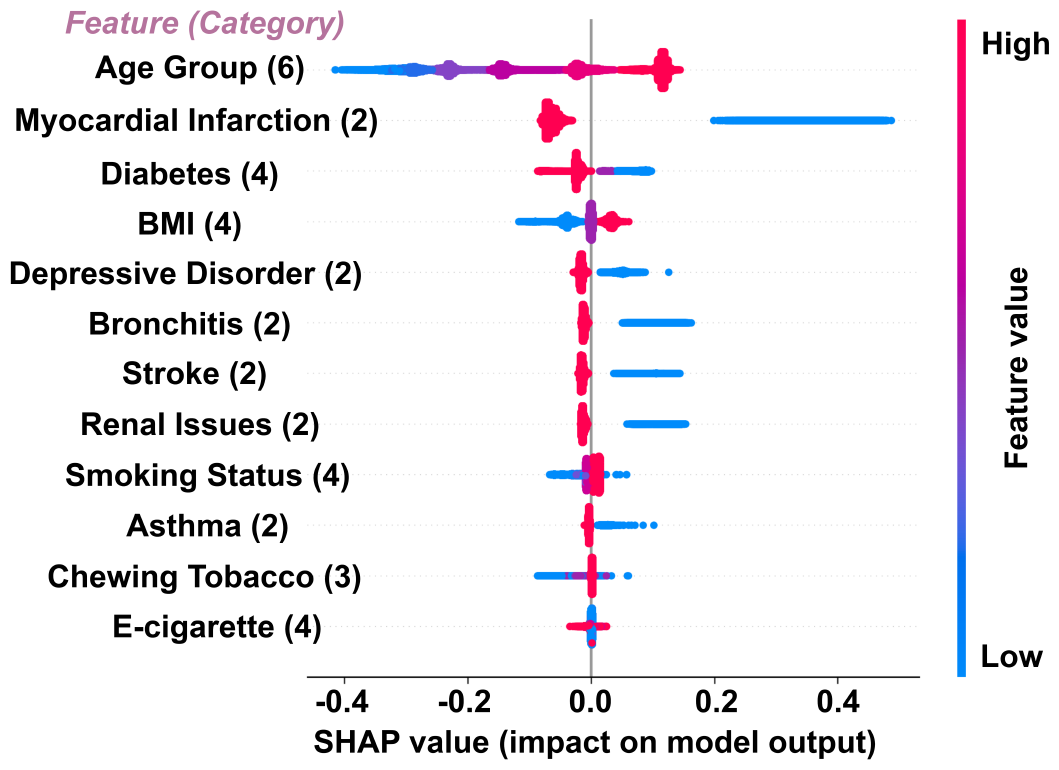
Feature effects on heart disease using the SHapley Additive exPlanations (SHAP). For the binary features, blue indicates individuals with a history of these diseases and red indicates individuals who have no issue in these regards. Age groups contain six categories (young adults, early adults, mid adults, late adults, mature adults, and elderly). The diabetes column contains four categories (diabetes, during pregnancy, no diabetes, and prediabetes). The BMI column contains four categories (underweight, normal weight, overweight, and obese). The smoking status contains four categories (chain smoker, irregular smoker, former, and never). The tobacco usage column contains three categories (regular, irregular, and no). The e-cigarette column contains four categories (never, regular, irregular, and former). The color hues vary from blue to purple to red for the input categories in multiclass features.

The impact of each category inside BMI, age group, and diabetes are shown in **Fig 7**. The elderly group has a greater contribution in predicting heart disease cases. Besides, elderly individuals with myocardial infarction (MI) contributed to predicting heart disease cases. Accordingly, the elderly with diabetes have a significant contribution in predicting heart disease cases. Similarly, females having diabetes during pregnancy have also contributed to predicting heart disease cases. Lastly, the obese condition in different age groups has a greater influence on predicting heart disease cases.

**Fig. 7.**
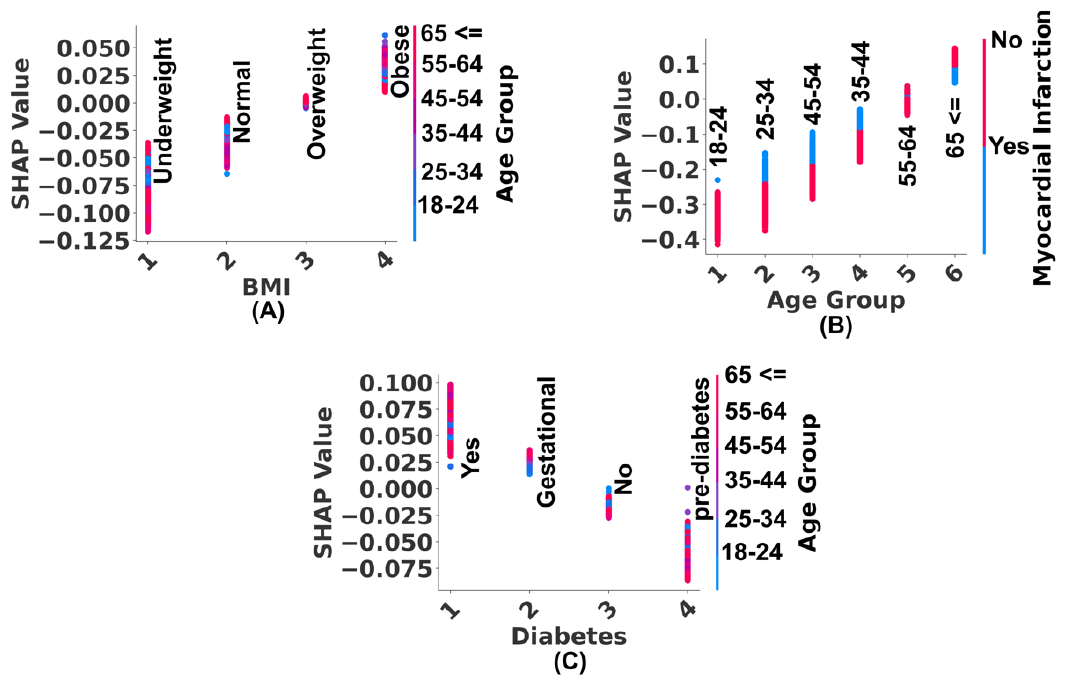
Effect of each category in multi-class features on heart disease using the SHapley Additive exPlanations (SHAP).

The impact of each category inside smoking, chewing tobacco, and e-cigarettes are shown in **Fig 8**. Young individuals with chain-smoking habits have a significant contribution in predicting heart disease cases. Accordingly, individuals with a habit of chewing tobacco have also contributed to predicting heart disease cases. Individuals with myocardial infarction in this category seem to have a minimal impact on the prediction of heart disease cases. Besides, Individuals with a habit of using e-cigarettes have an influence on the prediction of heart disease cases. The smoking transition column has been formed by performing feature engineering in the three columns including smoking status, chewing tobacco, and e-cigarettes. Lastly, the transition from cigarettes to chewing tobacco and e-cigarettes drastically reduces the prediction influence of heart disease cases among the elderly.

**Fig. 8.**
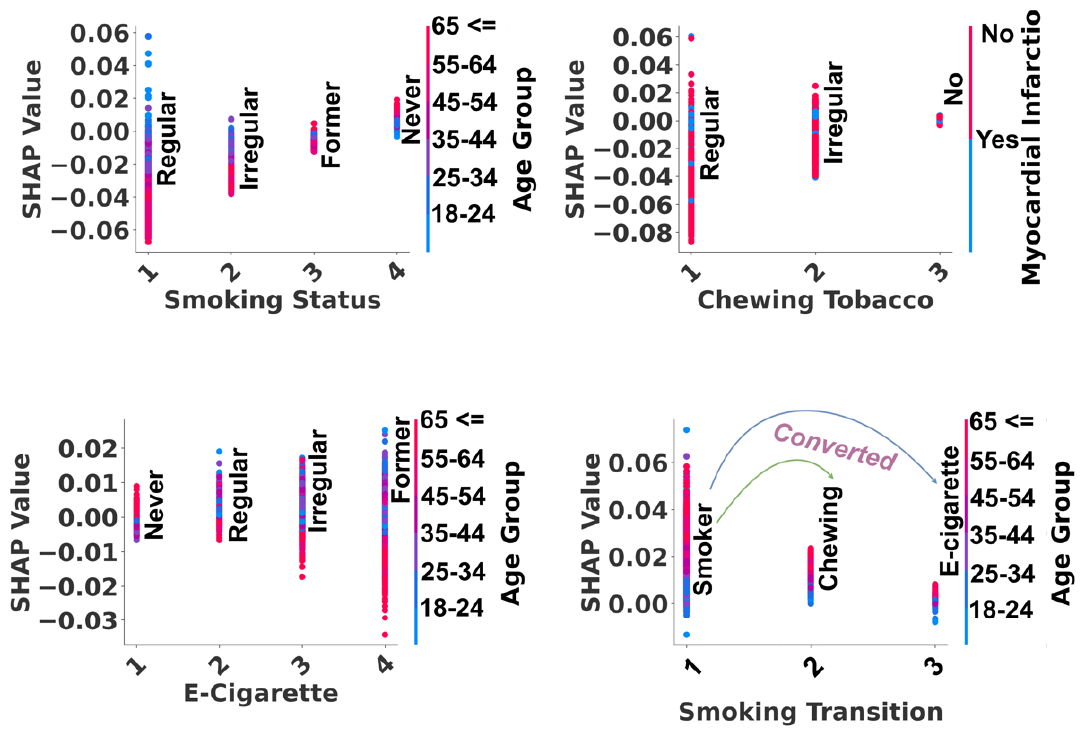
Effects of smoking, chewing tobacco, and e-cigarettes on heart disease using the SHapley Additive exPlanations (SHAP)

## IX. Discussion

The study has brought to light some important gaps in the body of research on the use of artificial intelligence (AI) in the prediction of heart disease. In the validation phases, the proposed GrowNet model has fluctuating outcomes. The training of the model was conducted on a synthetic balanced dataset. However, the test set was heavily unbalanced (172473 healthy and 6713 HD cases). So, the proposed model faced fluctuating outcomes when it introduced to heavily unbalanced train set. Srinivasan et al. [4] conducted a study that obtained above 0.97 of specificity and sensitivity but tested on very limited data, around 150 samples. Similarly, Chandrasekhar et al. [5] conducted a study that obtained an average recall of 0.97 but also tested on very limited data around 88 healthy and 96 HD records. However, Akkaya et al. [5] conducted a study employing SMOTE-Tomek Link to handle class imbalance. However, the obtained specificity (0.97) and sensitivity (0.27) were unbalanced when tested on a large dataset (51884 healthy and 4175 HD cases) [5]. In this way, the performance of the model can decrease when tested across real scenarios (unbalanced) [5] [6]. Nevertheless, the proposed GrowNet model obtained balanced specificity (0.74) and sensitivity (0.81) across a heavily unbalanced test set (172473 healthy and 6713 HD cases) using a Synthetic Refinement Pipeline to handle class imbalance. Besides, the proposed GrowNet model obtained comparatively balanced outcomes such as specificity (0.77, 0.87), and sensitivity (0.79, 0.77) across the other two versions of the dataset with 46640 and 369767 records respectively. The RUSBoost model seemed to have similar performance as the proposed GrowNet but the recall for the minority class (heart disease class) was comparatively lower (0.73) than the GrowNet model’s recall (0.81). The heart disease class is the most important in this study. Besides, the AUC score of the GrowNet (0.8709) was significantly better than the RUSBoost (0.7697). Similarly, the AUC score of ANN is almost the same as the GrowNet model but the recalls were heavily unbalanced (healthy: 0.64, HD: 0.88). Hence, the proposed GrowNet model obtained the most optimized and considerable outcomes compared to other models.

Additionally, risk factors for heart disease are identified by applying explainable AI (XAI) to the proposed GrowNet model. Individuals who have a history of heart attack, diabetes, obesity, depressive disorder, chronic bronchitis, stroke, renal issues, and asthma seemed to have a high SHAP [15] [16] probability score to predict heart disease cases. There is a link between these medical conditions and the heart. Hence, a person’s risk of developing heart disease can be raised by any of these conditions. Further, tobacco products including cigarettes, chewing tobacco, and e-cigarettes have an influence in the predicting of heart disease cases. These products can harm the condition of an individual’s heart. Hence, consuming these products regularly can be a significant risk factor for heart disease. Accordingly, individuals who started to use chewing tobacco or e-cigarettes instead of cigarettes seemed to have a significantly lower SHAP probability score than smokers. Hence, this can be stated that e-cigarettes or chewing tobacco have less influence on heart disease than cigarettes.

## Data Availability

https://www.cdc.gov/brfss/index.html

